# mBRSET: A Portable Retina Fundus Photos Benchmark Dataset for Clinical and Demographic Prediction

**DOI:** 10.1101/2024.07.11.24310293

**Authors:** Chenwei Wu, David Restrepo, Luis Filipe Nakayama, Lucas Zago Ribeiro, Zitao Shuai, Nathan Santos Barboza, Maria Luiza Vieira Sousa, Raul Dias Fitterman, Alexandre Durao Alves Pereira, Caio Vinicius Saito Regatieri, Jose Augusto Stuchi, Fernando Korn Malerbi, Rafael E. Andrade

## Abstract

*This paper introduces mBRSET, the first publicly available retina dataset captured using handheld retinal cameras in real-life, high-burden scenarios, comprising 5,164 images from 1,291 patients of diverse backgrounds. This dataset addresses the lack of ophthalmological data in low- and middle-income countries (LMICs) by providing a cost-effective and accessible solution for ocular screening and management. Portable retinal cameras enable applications outside traditional hospital settings, such as community health screenings and telemedicine consultations, thereby democratizing healthcare. Extensive metadata that are typically unavailable in other datasets, including age, sex, diabetes duration, treatments, and comorbidities, are also recorded. To validate the utility of mBRSET, state-of-the-art deep models, including ConvNeXt V2, Dino V2, and SwinV2, were trained for benchmarking, achieving high accuracy in clinical tasks diagnosing diabetic retinopathy, and macular edema; and in fairness tasks predicting education and insurance status. The mBRSET dataset serves as a resource for developing AI algorithms and investigating real-world applications, enhancing ophthalmological care in resource-constrained environments*.

## Background & Summary

Imaging exams play a pivotal role in diagnosing and monitoring ophthalmological pathologies^1^. Retinal fundus photos are widely implemented to register the ocular posterior segment, comprising the retina, optic disc, macula, and vessels, and are captured employing specialized cameras ^1^. However, there is a significant scarcity of ophthalmological data in low- and middle-income countries (LMICs), and the high costs of traditional imaging techniques further exacerbate this issue. Traditionally, retinal imaging has relied on tabletop fundus cameras, adhering to the guidelines outlined in the Early Treatment Diabetic Retinopathy Study ^2^. However, this approach poses accessibility challenges due to its cumbersome setup, spatial demands, intricate acquisition procedures, and substantial upfront investment, thus constraining its widespread adoption, which is particularly challenging in LMICs ^3,4^.

The emergence of compact, portable devices marks a significant stride toward a more accessible and cost-efficient screening process, particularly benefiting resource-limited regions, remote areas, and underrepresented communities ^3,5^. When coupled with timely intervention, these methods are crucial in preventing visual impairment ^5,6^. Moreover, the combination of artificial intelligence (AI) algorithms with portable devices has the potential to revolutionize medical care by streamlining screening, diagnosis, and monitoring processes, especially in resource-constrained settings ^7^. However, concerns regarding the accuracy and fairness of AI algorithms persist, primarily due to a lack of representative data and generalizable algorithms ^8^.

In low-and-middle-income countries (LMICs), the scarcity and uneven distribution of ophthalmologists relative to the population underscore the critical need for affordable and accessible ocular care ^9–12^. The use of portable retinal cameras not only reduces costs but also enables real-world applications outside of hospital settings, such as community health screenings and remote telemedicine consultations. Nevertheless, the existing ophthalmological datasets do not adequately reflect LMIC countries and this evolving retinal imaging modality ^13^.

To address these challenges, we present mBRSET, a groundbreaking advancement as the first publicly available diabetic retinopathy dataset composed of images captured with handheld retina cameras in real-life, high-burden scenarios, as shown in Table 1. The importance of the mBRSET dataset lies four-fold: First, by focusing on individuals from diverse ethnic backgrounds in Brazil, mBRSET addresses the underrepresentation of LMIC populations in current ophthalmological datasets. Second, mBRSET is the first publicly available dataset to feature images captured using handheld retina cameras, reflecting the growing trend of portable retinal imaging in resource-limited settings. Third, the dataset is collected in authentic, high-burden clinical settings, ensuring its relevance to real-world ocular screening and management challenges. Fourth, by including demographic information such as sex, education level, and insurance status, mBRSET enables researchers to assess the fairness and generalizability of AI algorithms across different subpopulations. Finally, the dataset serves as a valuable resource for developing and validating AI algorithms that can streamline the screening, diagnosis, and monitoring of diabetic retinopathy and other ocular pathologies in resource-constrained environments.

By providing a comprehensive and representative dataset, mBRSET has the potential to drive advancements in accessible ocular care, particularly in LMICs, and contribute to the development of fair and accurate AI-assisted diagnostic tools for the early detection and management of vision-threatening conditions.

## Methods

The work received approval from the Institutional Review Board of Instituto de Ensino Superior Presidente Tancredo de Almeida Neves (IPTAN) under protocol number CAAE 64219922.3.0000.9667. It encompassed retinal fundus photos alongside clinical and demographic data. Notably, all identifiable patient information was removed from the images within this dataset to ensure confidentiality and adherence to ethical standards. All patients have given written consent to the image capture and open publication.

### Data sources

We gathered data from participants attending the Itabuna Diabetes Campaign, held in November 2022 in Itabuna, Bahia State, located in Northeast Brazil. Bahia State boasts a rich tapestry of ethnicities, with approximately half of its population tracing their lineage to European ancestry, around 40% with roots in African heritage, and 10% carrying Native American ancestry ^14^. This mosaic of ancestries contributes to the diverse fabric of society in Bahia, showcasing the fusion of cultures unique to the region.

The annual Itabuna Diabetes Campaign is a pivotal event, galvanizing numerous city residents to participate in activities focused on diabetes awareness, screening, and treatment of diabetes-related complications ^15^. Upon consenting to participate, individuals completed a questionnaire detailing demographic information and self-reported clinical characteristics before undergoing ocular imaging.

### Data acquisition

The images contained within this dataset were acquired using the Phelcom Eyer (Phelcom Technologies, Sao Carlos, Brazil), a portable, handheld retinal fundus camera that integrates with a Samsung Galaxy S10 running Android 11. Operating at a 45-degree angle, this camera employs a high-resolution 12-megapixel sensor to generate images with dimensions of 1600x1600 pixels. Notably, the Eyer camera boasts autofocus control capabilities, spanning a diopter range from -20 to +20.

Trained personnel proficient in the operation of the Eyer camera conducted the acquisition of retinal photos after pharmacological mydriasis. Furthermore, pertinent demographic details and relevant medical information were gathered through structured patient interviews, ensuring a comprehensive dataset for analysis.

### Dataset preparation

Every fundus photo within the dataset was removed of all identifying information, including file identification and sensitive data such as patient names. Each image underwent thorough scrutiny to confirm the absence of any protected health information. Subsequently, the images were extracted from the Eyer cloud system in JPEG format without undergoing any preprocessing techniques.

It’s important to note that the images within the dataset may feature a viewpoint centered either on the macula or the optic disc. However, the dataset excludes fluorescein angiogram photos and any non-retinal images, ensuring a focused and relevant collection for analysis.

### Labeling

Two ophthalmologists, specialized in retina and vitreous conditions, annotated all images based on predefined criteria established by the research group ^16^. In instances of disagreement regarding the grading of diabetic retinopathy, a third senior specialist provided adjudication. To ensure a robust evaluation of inter-rater reliability, Kappa and weighted kappa statistics were employed to assess agreement between graders.

The labeled characteristics encompassed

Quality control parameters: This evaluation included identifying artifacts, such as issues with image focus, illumination disparities, and the presence of dust. Each image was then categorized based on its quality, determining whether it was deemed satisfactory or unsatisfactory for clinical assessment.

Diabetic retinopathy classification: The classifications of diabetic retinopathy and diabetic macular edema were determined using the International Clinical Diabetic Retinopathy (ICDR) grading system 17. (See **Table 1** for details.)

**Table 1:** International Clinic Diabetic Retinopathy (ICDR) grading.

| Classification | International Classification of Diabetic Retinopathy <sup>17</sup> |
| --- | --- |
| 0 - Normal | No abnormalities |
| 1 - Mild non-proliferative DR | Microaneurysms only |
| 2- Moderate non-proliferative DR | More than just microaneurysms but less than severe non-proliferative diabetic retinopathy |
| 3 - Severe non-proliferative DR | Any of the following:<br>> 20 intra-retinal hemorrhages in each of 4 quadrants,<br>definite venous beading in $\geq 2$ quadrants,<br>prominent intraretinal microvascular abnormalities in $\geq 1$ quadrant, or no signs of proliferative retinopathy |
| 4 - Proliferative DR | One or more of the following: neovascularization and/or vitreous or preretinal hemorrhages and/or panphotocoagulation scars |
| Macular edema | Exudates or apparent thickening within one disc diameter from the fovea |

## Data Records

The dataset comprises comprehensive records of diabetic patients, collected during the Itabuna Diabetes Campaign. The table below summarizes the data fields available, their descriptions, and the coding used for categorical variables. The dataset is composed of a file containing metadata with labels and demographic information of the patient in a csv format, and a directory with the corresponding images in a .jpg format. The images are identified by a unique id used as filename, and are linked with the metadata csv file using the column “file”.

The csv dataset includes basic demographic information such as age, sex, and duration of diabetes diagnosis (age, sex, dm_time). Clinical information is detailed, capturing treatment and comorbid conditions, including insulin use (insulin, insulin_time), oral diabetes medication (oraltreatment_dm), hypertension (systemic_hypertension), and other diabetes-related complications (vascular_disease, acute_myocardial_infarction, nephropathy, neuropathy, diabetic_foot).

Lifestyle factors such as smoking status (smoking), alcohol consumption (alcohol consumption), and obesity (obesity) are recorded. Additionally, healthcare and education details are included, such as insurance status (insurance) and educational attainment (educational_level).

The table also contains detailed information on the retinal images, including the image identifier (file), laterality (laterality), presence of artifacts (final_artifacts), image quality assessment (final_quality), and the ICDR score (final_icdr) for retinopathy. The presence of macular edema (final_edema) is also recorded.

This detailed structure ensures that the dataset is comprehensive and provides valuable insights into the patient population and the quality of the retinal images collected.

**Table 1.** Data Records Overview.

| Field | Description | Values |
| --- | --- | --- |
| file | Image identifier | String |
| patient | Patient identifier | Integer |
| age | Age of the patient in years | Integer |
| sex | Gender of the patient | 0 = Female, 1 = Male |
| dm_time | Duration of diabetes diagnosis in years | Integer |
| insulin | Use of insulin | 0 = No, 1 = Yes |
| insulin_time | Duration of insulin use in years | Integer |
| oraltreatment_dm | Use of oral drugs for diabetes | 0 = No, 1 = Yes |
| systemic_hypertension | Diagnosis of systemic arterial hypertension | 0 = No, 1 = Yes |
| insurance | Health insurance status | 0 = No, 1 = Yes |
| educational_level | Highest educational level achieved | 1 = Illiterate,<br>2 = Incomplete Primary,<br>3 = Complete Primary,<br>4 = Incomplete Secondary,<br>5 = Complete Secondary,<br>6 = Incomplete Tertiary,<br>7 = Complete Tertiary |
| alcohol_consumption | Regular alcohol consumption | 0 = No, 1 = Yes |
| smoking | Active smoking status | 0 = No, 1 = Yes |
| obesity | Diagnosis of obesity | 0 = No, 1 = Yes |
| vascular_disease | Diabetes-related vascular disease | 0 = No, 1 = Yes |
| acute_myocardial_infarction | History of acute myocardial infarction | 0 = No, 1 = Yes |
| nephropathy | Diabetes-related nephropathy | 0 = No, 1 = Yes |
| diabetic_foot | Presence of diabetic foot | 0 = No, 1 = Yes |
| laterality | Laterality of retinal fundus photo | Left, Right |
| final_artifacts | Presence of artifacts in the retinal image | Yes, No |
| final_quality | Final quality assessment of the retinal image | Yes, No |
| final_icdr | ICDR score for retinopathy assessment | 0=No retinopathy,<br>1=Mild non-proliferative,<br>2=Moderate non-proliferative,<br>3=Severe non-proliferative,<br>4=Proliferative diabetic retinopathy and post-laser status |
| final_edema | Presence of macular edema | Yes, No |

### Patient level analysis

#### Demographic Analysis

The dataset comprises 5,164 images obtained from a cohort of 1,291 patients. Among these patients, 451 (34.93%) are male, while 840 (65.06%) are female. The average age of the participants is 61.44 years, with a standard deviation of 11.63 years. **Table 2** provides a detailed breakdown of patient demographics, including insurance status and educational levels.

**Table 2:** Patient’s demographics characteristics.

| Variable | Missing |  | Num. Patients | Num. Study |
| --- | --- | --- | --- | --- |
| Patients, n | 0 |  | 1291 | 5164 |
| sex, n (%) | 0 | Female | 840 (65.1) | 3360 |
|  |  | Male | 451 (34.9) | 1804 |
| insurance, n (%) | 12 | No | 1180 (92.3) | 4720 |
|  |  | Yes | 99 (7.7) | 396 |
| educational_level, n (%) |  | Illiterate (1) | 176 (13.8) | 704 |
|  |  | Incomplete Primary (2) | 539 (42.2) | 2156 |
|  |  | Complete Primary (3) | 116 (9.1) | 464 |
|  |  | Incomplete Secondary (4) | 120 (9.4) | 480 |
|  |  | Complete Secondary (5) | 247 (19.3) | 988 |
|  |  | Incomplete Tertiary (6) | 21 (1.6) | 84 |
|  |  | Complete Tertiary (7) | 59 (4.6) | 236 |
| age, mean (SD) | 0 |  | 61.4<br>(11.6) | 61.45(11.63) |

The demographic analysis of the mBRSET dataset reveals a diverse cohort of patients, with a majority being female (65.06%). The mean age of the participants is 61.44 years, reflecting a predominantly elderly population, which is typical for studies focused on diabetes-related ocular conditions. A significant proportion of the cohort lacks health insurance (92.3%), highlighting potential socio-economic challenges faced by the participants. Educational attainment is varied, with the largest group having incomplete primary education (42.2%). These demographic insights underscore the representativeness and relevance of the mBRSET dataset in studying diabetic retinopathy and related conditions in diverse, real-world settings.

#### Clinical Analysis

**Table 3** provides detailed statistics on the clinical characteristics of the patients, including the prevalence of comorbid conditions and lifestyle factors. The clinical characteristics of the mBRSET dataset reveal significant insights into the health status and comorbidities of the patient cohort. The average duration of diabetes among participants is approximately 9.53 years, indicating a substantial history of the disease within this population. A majority of the patients (84.61%) are undergoing oral treatment for diabetes, while a smaller proportion (20.79%) are on insulin therapy.

**Table 3:** Patients’ clinical characteristics.

| Variable | Missing |  | Num. Patients | Num. Study |
| --- | --- | --- | --- | --- |
| n |  |  | 1291 | 5164 |
| systemic_hypertension, n (%) | 11 | No | 366 (28.6) | 1464 |
|  |  | Yes | 914 (71.4) | 3656 |
| alcohol_consumption, n (%) | 19 | No | 1094 (86.0) | 4376 |
|  |  | Yes | 178 (14.0) | 712 |
| smoking, n (%) | 22 | No | 1188 (93.6) | 4752 |
|  |  | Yes | 81 (6.4) | 324 |
| obesity, n (%) | 19 | No | 1170 (92.0) | 4680 |
|  |  | Yes | 102 (8.0) | 408 |
| vascular_disease, n (%) | 19 | No | 1055 (82.9) | 4220 |
|  |  | Yes | 217 (17.1) | 868 |
| acute_myocardial_infarction, n (%) | 21 | No | 1172 (92.3) | 4688 |
|  |  | Yes | 98 (7.7) | 392 |
| nephropathy, n (%) | 20 | No | 1225 (96.4) | 4900 |
|  |  | Yes | 46 (3.6) | 184 |
| neuropathy, n (%) | 19 | No | 1217 (95.7) | 4868 |
|  |  | Yes | 55 (4.3) | 220 |
| diabetic_foot, n (%) | 27 | No | 1091 (86.3) | 4364 |
|  |  | Yes | 173 (13.7) | 692 |

Systemic hypertension is prevalent among the cohort, with 71.4% of patients diagnosed with this condition. Alcohol consumption and smoking are less common, reported by 14.0% and 6.4% of the participants, respectively. Obesity is identified in 8.0% of the patients, and vascular disease is present in 17.1%.

The incidence of acute myocardial infarction stands at 7.7%, while nephropathy and neuropathy are reported by 3.6% and 4.3% of the patients, respectively. Additionally, 13.7% of the participants have diabetic foot. These clinical characteristics highlight the complex and multifaceted health challenges faced by the diabetic population in the mBRSET dataset, underscoring the importance of comprehensive care and monitoring for these individuals.

### Image analysis

In terms of image findings within the dataset, a comprehensive analysis revealed that among the total of 4,885 images assessed, 3,759 images (76.79%) exhibited no indications of diabetic retinopathy (DR). Mild non-proliferative DR was identified in 272 images (5.56%), while moderate non-proliferative DR was present in 570 images (11.64%). Furthermore, 82 images (1.67%) exhibited severe non-proliferative DR, and 212 images (4.33%) displayed proliferative DR (**Figure 1**). Additionally, macular edema was observed in 427 images (8.69%). (See **Table 4** for a comprehensive breakdown).

**Figure 1:**
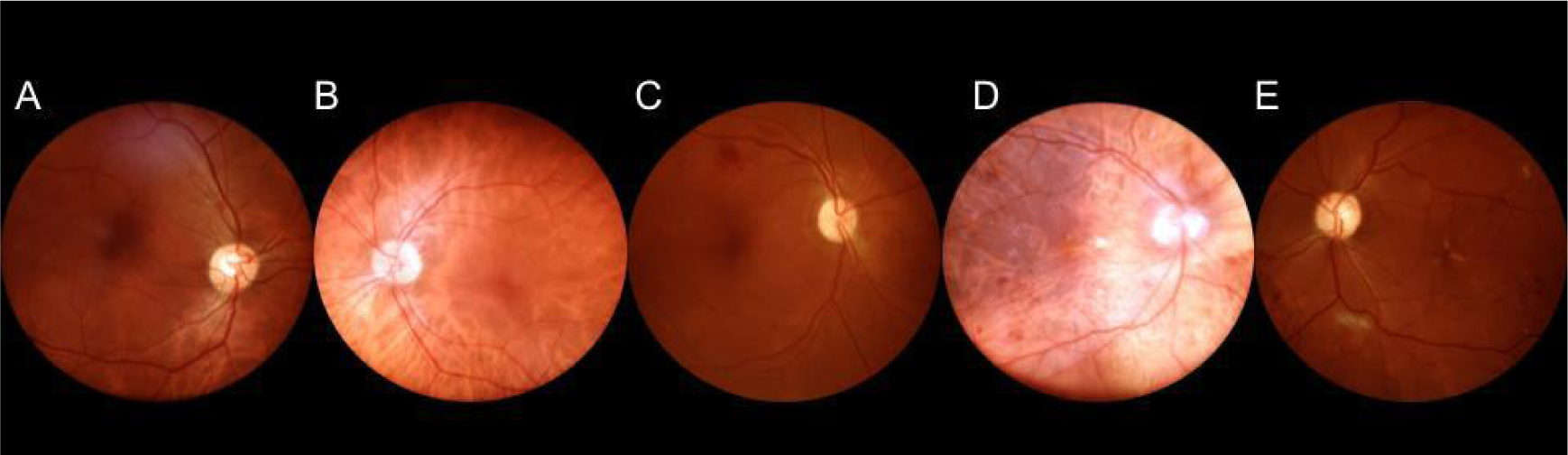
Diabetic Retinopathy images from mBRSET. A: Normal retina, B: Mild non-proliferative DR, C: Moderate non-proliferative DR, D: Severe non-proliferative DR, E: Proliferative DR.

**Table 4:**
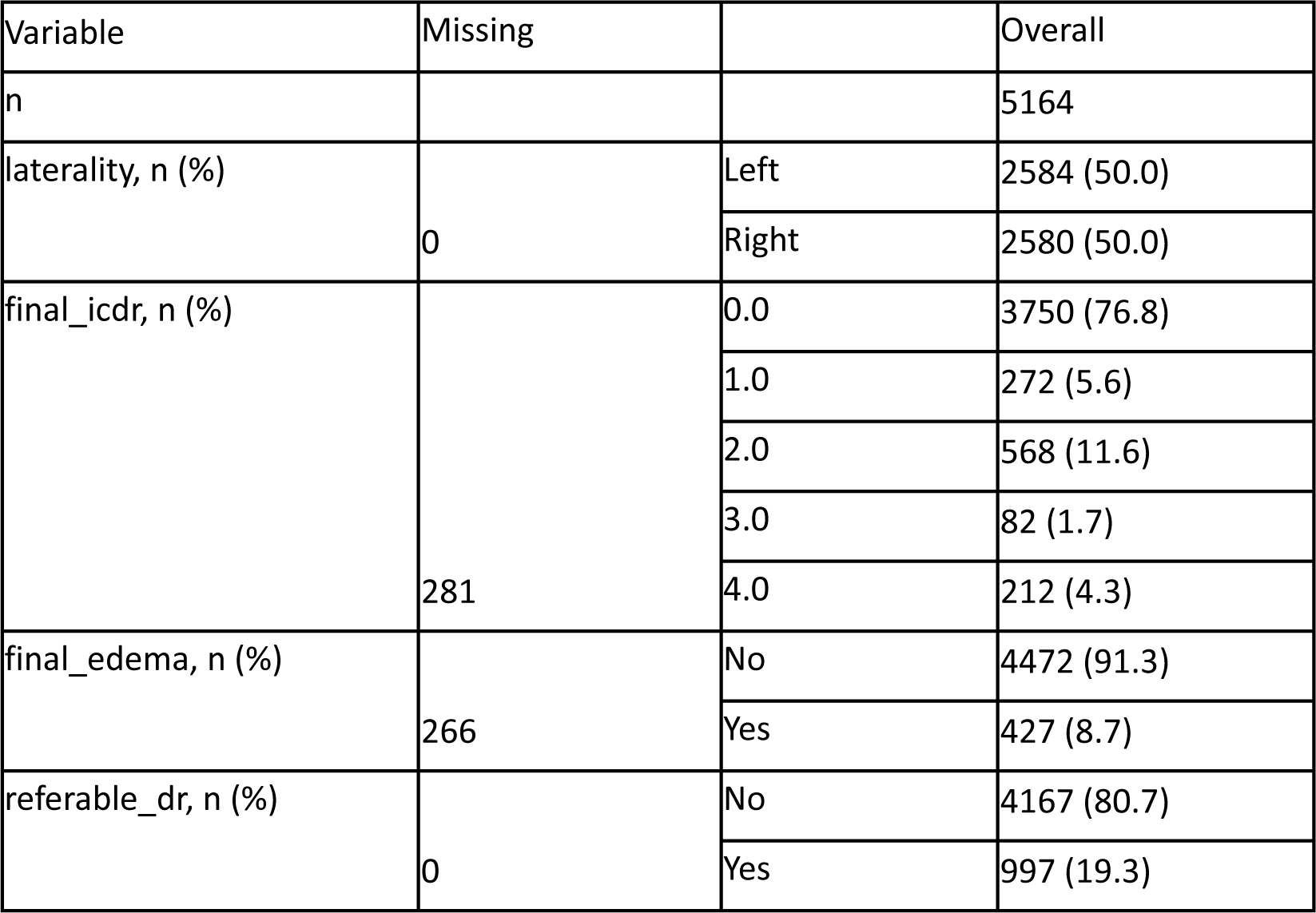
Image level analysis.

| Variable | Missing |  | Overall |
| --- | --- | --- | --- |
| n |  |  | 5164 |
| laterality, n (%) | 0 | Left | 2584 (50.0) |
|  |  | Right | 2580 (50.0) |
| final_icdr, n (%) | 281 | 0.0 | 3750 (76.8) |
|  |  | 1.0 | 272 (5.6) |
|  |  | 2.0 | 568 (11.6) |
|  |  | 3.0 | 82 (1.7) |
|  |  | 4.0 | 212 (4.3) |
| final_edema, n (%) | 266 | No | 4472 (91.3) |
|  |  | Yes | 427 (8.7) |
| referable_dr, n (%) | 0 | No | 4167 (80.7) |
|  |  | Yes | 997 (19.3) |

Regarding inter-grader agreement, the Weighted Kappa for the ICDR score was calculated at 0.863, indicating near-perfect agreement among the graders in assessing diabetic retinopathy severity. For the identification of macular edema, the Weighted Kappa stood at 0.618, indicating substantial agreement between the graders.

In terms of quality distribution, it’s noted that 4,283 images (82.7%) exhibit some form of artefact, while 4,833 images (94.3%) boast good quality, facilitating effective image assessment (**Figure 2**). (Refer to **Table 5** for a detailed breakdown.)

**Figure 2:**
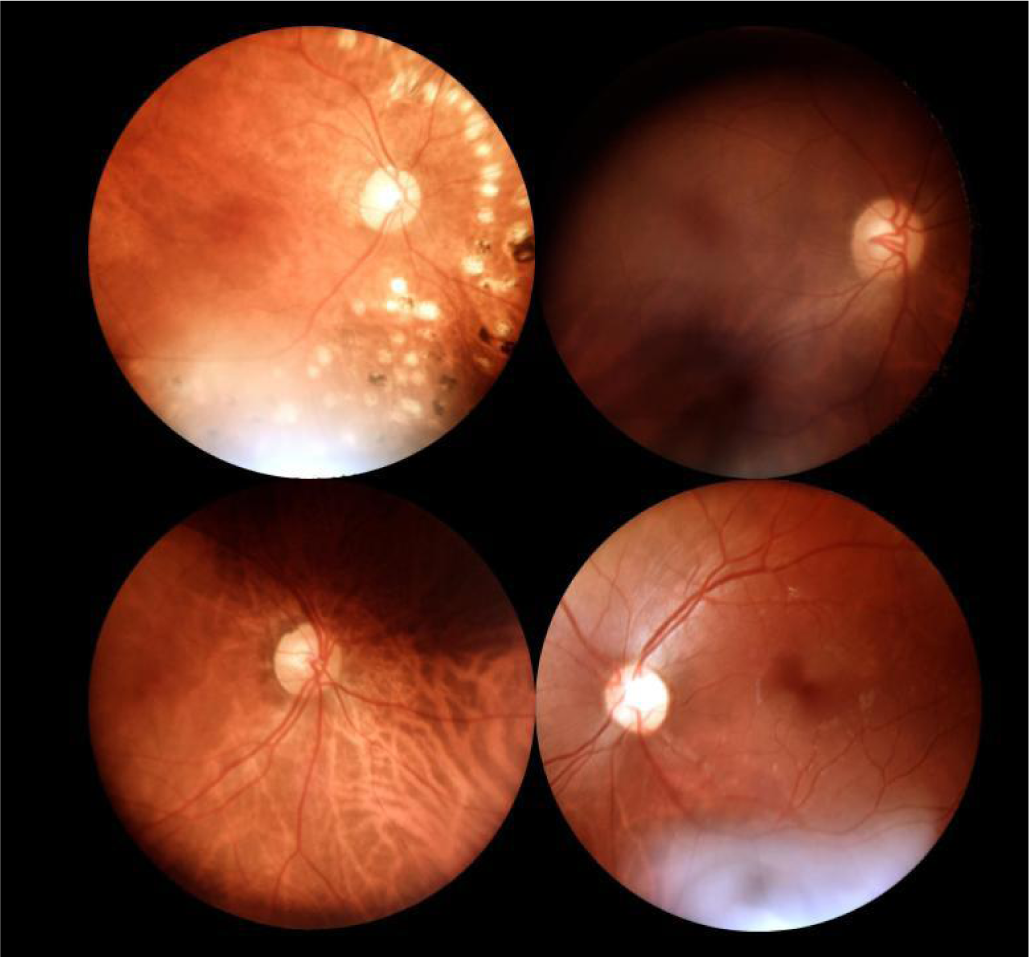
Images with artefacts from mBRSET - illumination and focus.

**Table 5:** Image artefacts and quality distribution.

| Variable | Missing |  | Overall |
| --- | --- | --- | --- |
| n |  |  | 5164 |
| final_artifacts, n (%) | 0 | No | 892 (17.3) |
|  |  | Yes | 4272 (82.7) |
| final_quality, n (%) | 0 | No | 292 (5.7) |
|  |  | Yes | 4872 (94.3) |

## Technical Validation

To demonstrate the utility and validate the quality of the mBRSET dataset, we present several use cases and benchmarks for future research. The code required to reproduce these experiments is available in a public GitHub repository (see Code Availability section).

### Dataset Comparison

To provide a comprehensive evaluation, we compared mBRSET with other publicly available retinal fundus datasets, such as BRSET ^18^, EyePACS ^19^, Messidor 2 ^20^, and the Retinal Fundus Multi-Disease Image Dataset (RFMiD) ^21^. This comparison focused on key attributes such as view position, labels, dataset size, availability of clinical and demographic information, diabetic retinopathy classification, nationality, and camera type (see **Table 6**).

**Table 6:**
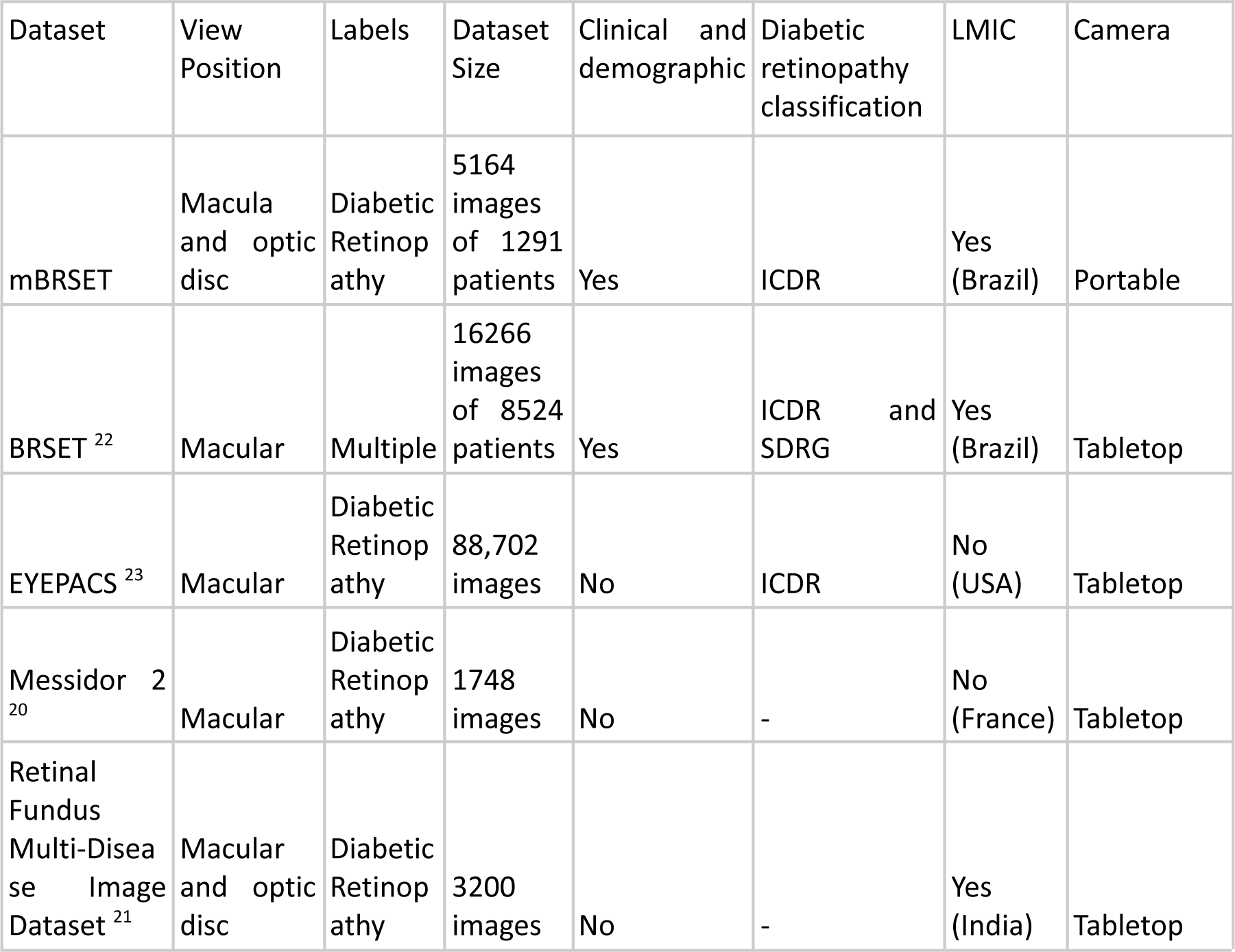
Comparison of new dataset (mBRSET), BRSET, and publicly available datasets.

| Dataset | View Position | Labels | Dataset Size | Clinical and demographic | Diabetic retinopathy classification | LMIC | Camera |
| --- | --- | --- | --- | --- | --- | --- | --- |
| mBRSET | Macula and optic disc | Diabetic Retinopathy | 5164 images of 1291 patients | Yes | ICDR | Yes (Brazil) | Portable |
| BRSET<sup>22</sup> | Macular | Multiple | 16266 images of 8524 patients | Yes | ICDR and SDRG | Yes (Brazil) | Tabletop |
| EYEPACS<sup>23</sup> | Macular | Diabetic Retinopathy | 88,702 images | No | ICDR | No (USA) | Tabletop |
| Messidor 2<sup>20</sup> | Macular | Diabetic Retinopathy | 1748 images | No | - | No (France) | Tabletop |
| Retinal Fundus Multi-Disease Image Dataset<sup>21</sup> | Macular and optic disc | Diabetic Retinopathy | 3200 images | No | - | Yes (India) | Tabletop |

### Prediction Tasks

We validate the mBRSET dataset by applying state-of-the-art deep learning models to several clinically relevant prediction tasks. As diabetic retinopathy is a leading cause of preventable blindness among adults and the most explored disease in public retinal image datasets ^24–26^, we focus on diabetic retinopathy prediction and clinical diagnosis of macular edema. Specifically, we conduct experiments on binary classification (Healthy vs Diabetic Retinopathy), three-class classification (Normal ICDR 0, Non-proliferative ICDR 1-3, and Proliferative Retinopathy ICDR 4), and binary macular edema prediction. Additionally, to explore the potential for fairness analysis using demographic information, we include prediction tasks for sex, educational level (Illiterate vs Educated), and insurance status (Has insurance vs No) ^27,28^.

### Network Frameworks

As shown in Figure 3, we employ ConvNext V2, DINO V2, and Swin V2 for benchmarking purposes, which represent state-of-the-art convolutional neural networks (CNNs) and vision transformers (ViTs), respectively.

**Figure 3.**
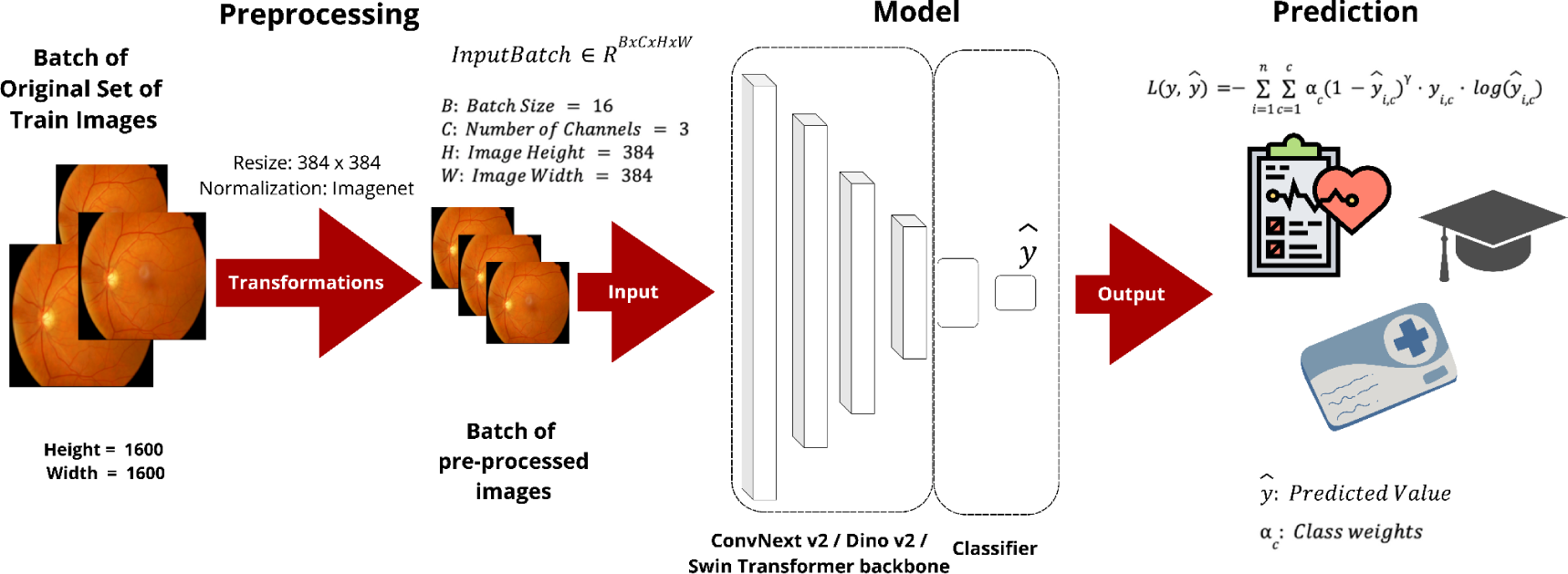
Benchmarking of State-of-the-art Deep Learning Models.

ConvNext V2 is an improved version of the ConvNext architecture, designed to achieve high performance while maintaining computational efficiency ^29,30^. It incorporates Masked Auto-Encoder (MAE) pre-training, depthwise convolutions, inverted bottlenecks, large kernel sizes, and regularization techniques. These enhancements help capture complex features and improve performance on tasks requiring a global understanding of the image.

DINO V2, an improved version of the DINO architecture, is a state-of-the-art vision transformer that learns self-supervised visual representations ^31^. It leverages self-supervised pre-training on large-scale datasets, a multi-crop strategy, attention mechanisms, and scalability to learn rich and generalizable visual features. These architectures offer advantages over their predecessors, such as ResNets and the original ViT, which have been widely used in prior retinal imaging research ^32–34^.

SwinV2 is another state-of-the-art vision transformer that builds upon the success of the original Swin Transformer ^35^. It introduces several architectural improvements, such as the use of a hierarchical structure, shifted window attention, and cross-window connections. These designs enable SwinV2 to capture both local and global contexts, leading to improved performance on a wide range of vision tasks.

#### Data Preprocessing

The mBRSET dataset was divided into training (70%), validation (10%), and testing sets (20%) using all the images available in the dataset. We used a grouped stratification strategy to ensure a representative distribution of classes and no patient data leakage across different sets. The images in the dataset present variations in height, ranging from 874 to 2304 pixels, and width, ranging from 951 to 2984 pixels. To ensure consistency with the model’s input, all the images were transformed into the same resolution. In the main experiments, the images are resized to 384*384 and normalized by dividing them into 255 to ensure all pixel values are between 0 and 1. Also, all the images were normalized using imagenet normalization statistics.

### Training and Evaluation Details

Models are initialized from ImageNet pre-trained weights available on Huggingface, and finetuned on the full training set for a maximum of 50 epochs, with early stopping based on F1 score improvements on the validation set (patience: 10 epochs) to prevent overfitting. The Adam optimizer is employed with learning rates of 1e-5 and weight decay of 1e-6 for all tasks. We used a batch size of 16 and one single Nvidia A40 GPU with 48 GB VRAM for all experiments.

To address the class imbalance issue, we use focal loss with inverse class weights [2]. The focal loss function focuses on hard-to-classify examples by down-weighting the loss assigned to well-classified examples, thus addressing the class imbalance more effectively. The focal loss is defined by the parameters α and γ, where α is the weighting factor for each class, and γ (set to 2) is the focusing parameter that reduces the relative loss for well-classified examples. The focal loss function can be expressed as in equation (1):

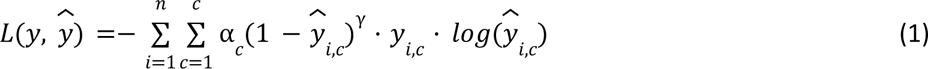

where *c* represents the number of classes, *n* represents the batch size, α_*c*_ represents the weights of class *c*, *y*_*i*,*c*_ is the ground truth, and 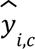 is the predicted value for class *c* for the i-th sample.

Accuracy, F1, precision, and recall score metrics were also used to assess model performance under class imbalance. The F1 score, which can be seen in equation 2, is a harmonic mean of precision and recall. For the multiclass problems, the macro average was applied to the F1 score to avoid biased results. Full results are shown in Table 7.

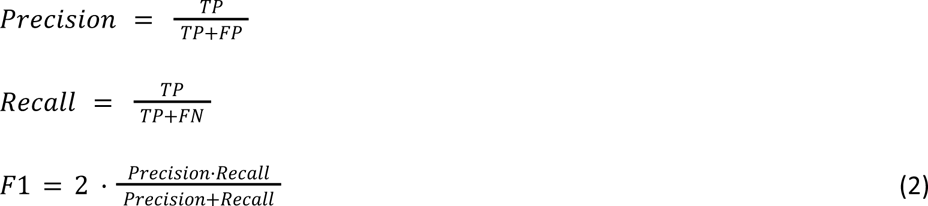

**Table 7:** Test Performance of Deep Learning Models Across Tasks.

| Task | Model | Accuracy | F1-score | Precision | Recall |
| --- | --- | --- | --- | --- | --- |
| Diabetic Retinopathy (3-class) | Swin-V2 Large <sup>35</sup> | 89.06 | 80.26 | 82.27 | 78.52 |
|  | DINO-V2 Large <sup>31</sup> | 87.51 | 79.19 | 78.64 | 79.92 |
|  | ConvNext-V2 Large <sup>29</sup> | 88.67 | 80.33 | 83.99 | 77.48 |
| Diabetic Retinopathy (binary) | Swin-V2 Large <sup>35</sup> | 89.55 | 85.98 | 89.07 | 83.89 |
|  | DINO-V2 Large <sup>31</sup> | 90.03 | 87.4 | 87.61 | 87.19 |
|  | ConvNext-V2 Large <sup>29</sup> | 89.55 | 86.26 | 88.23 | 84.77 |
| Macular Edema | Swin-V2 Large <sup>35</sup> | 92.55 | 82.66 | 85.54 | 80.38 |
|  | DINO-V2 Large <sup>31</sup> | 92.64 | 82.94 | 85.69 | 80.74 |
|  | ConvNext-V2<br>Large <sup>29</sup> | 92.64 | 83.06 | 85.51 | 81.04 |
| Insurance<br>Prediction | Swin-V2<br>Large <sup>35</sup> | 91.38 | 76.11 | 73.36 | 80.02 |
|  | DINO-V2<br>Large <sup>31</sup> | 90.22 | 71.4 | 69.9 | 73.28 |
|  | ConvNext-V2<br>Large <sup>29</sup> | 89.84 | 71.81 | 69.53 | 75.1 |
| Education<br>Prediction | Swin-V2<br>Large <sup>35</sup> | 72.73 | 72.33 | 72.34 | 72.33 |
|  | DINO-V2<br>Large <sup>31</sup> | 70.48 | 69.94 | 70.04 | 69.87 |
|  | ConvNext-V2<br>Large <sup>29</sup> | 70.77 | 70.18 | 70.35 | 70.09 |
| Gender<br>Prediction | Swin-V2<br>Large <sup>35</sup> | 83.25 | 81.14 | 82.06 | 80.46 |
|  | DINO-V2<br>Large <sup>31</sup> | 82.89 | 80.83 | 81.51 | 80.30 |
|  | ConvNext-V2<br>Large <sup>29</sup> | 85.67 | 84.38 | 84.11 | 84.69 |

### Clinical Diagnosis Results

Diabetic Retinopathy Classification: For the 3-class classification task, the models achieved F1 scores ranging from 79.19 to 80.33. ConvNext-V2 Large obtained the highest F1 score of 80.33, followed closely by Swin-V2 Large with 80.26.

In the binary classification setting, DINO-V2 Large outperformed the other models, achieving an F1 score of 87.4. While the accuracy scores were generally high (89.55% to 90.03%), the F1 scores provide a more reliable assessment of the model’s performance, considering the class imbalance in the dataset.

Macular Edema Detection: All three models demonstrated strong performance in detecting macular edema, with F1 scores above 82%. ConvNext-V2 Large achieved the highest F1 score of 83.06, closely followed by DINO-V2 Large (82.94) and Swin-V2 Large (82.66). The accuracy scores for macular edema detection were consistently high, ranging from 92.55% to 92.64%.

These results demonstrate that the mBRSET dataset can provide diagnostic performance on par with datasets obtained using traditional table-top cameras.

### Demographics Predictions Results

Gender Prediction: The models achieved F1 scores between 80.83 and 84.38 for gender prediction. ConvNext-V2 Large obtained the highest F1-score of 84.38, demonstrating its ability to capture gender-related features from the retinal images. The accuracy scores for gender prediction ranged from 82.89% to 85.67%. These results are consistent with previous works that indicate retinal fundus photos contain gender-specific information and can be inferred by deep computer vision models ^27^.

Education Prediction: The models’ performance in predicting education level was relatively lower compared to other tasks, with F1-scores ranging from 69.94 to 72.33. Swin-V2 Large achieved the highest F1-score of 72.33, indicating the challenging nature of inferring education status from retinal images. The accuracy scores for education prediction varied between 70.48% and 72.73%.

Insurance Prediction: The models obtained F1 scores ranging from 71.4 to 76.11 for insurance status prediction. Swin-V2 Large outperformed the other models with an F1 score of 76.11. The accuracy scores for insurance prediction were higher, ranging from 89.84% to 91.38%, but the F1 scores provide a more balanced assessment considering the class imbalance.

The demographic prediction results reveal previously unexplored patterns and contribute to our understanding of the complex interplay between socioeconomic factors and health outcomes. The deep learning models’ ability to infer demographic and socioeconomic information from retinal images highlights the potential of this approach for studying population health and addressing healthcare inequalities. Although the F1 scores for education and insurance prediction were lower than 80%, the fact that the deep networks were able to learn associations between socioeconomic disparities and retinal images is a significant finding. Previous statistical analyses have discussed the relationship between socioeconomic factors and retinal health ^36–39^, but this is the first time that deep learning models have successfully demonstrated this connection. These results open up new avenues for research and emphasize the importance of considering socioeconomic factors in the development and application of AI-assisted diagnostic tools. By leveraging the information captured in retinal images, we can gain valuable insights into population health and work towards more equitable healthcare solutions.

These results indicate that the mBRSET dataset, coupled with advanced deep learning models like ConvNeXt-V2, can effectively support various clinical predictions and demographic analyses on par with traditional tabletop cameras. The high accuracy and robust performance metrics demonstrate the dataset’s reliability and quality.

## Usage Notes

*The mBRSET dataset is publicly accessible through **Physionet**. This dataset comprises a comprehensive collection of images, along with detailed metadata and labels. To facilitate effective utilization of the dataset, we have provided a CSV file that contains essential information for each image. The CSV file includes a ‘file’ column, which lists the ID of each image without the extension (.jpg). Researchers can use this column to correlate the metadata and labels with the corresponding images*.

*Our GitHub repository is designed to support researchers in leveraging the mBRSET dataset effectively. It includes:*

- *Python Code for Computer Vision Models: We provide implementations of over 20 state-of-the-art computer vision models using PyTorch. This includes models specifically employed in this paper, ensuring that researchers can replicate our experiments or extend them using different models*.
- *Helper Functions: To streamline the preprocessing and experimental workflow, our repository includes a suite of helper functions. These functions cover a range of tasks from data preprocessing to model evaluation, facilitating a seamless research experience*.
- *Jupyter Notebooks: For ease of use and reproducibility, we have included Jupyter notebooks containing the code for the experiments conducted in this paper. These notebooks provide a detailed step-by-step guide, allowing researchers to understand the experimental setup, run the experiments, and visualize the results*.

### Accessing the mBRSET Dataset and Resources

- *Physionet Dataset: The mBRSET dataset can be accessed through Physionet at the following link (*https://www.physionet.org/content/mbrset/1.0/)
- *GitHub Repository: Our repository, which includes the Python code, helper functions, and Jupyter notebooks, is available here:* https://github.com/luisnakayama/mBRSET

### Example of Using the CSV File for Metadata and Labels

*The CSV file provided with the dataset contains a ‘file’ column, which is crucial for accessing the metadata and labels associated with each image. For example, if an image file is named 00123.jpg, the ‘file’ column will contain 00123. Researchers can use this identifier to retrieve all relevant information for that image, streamlining the process of data handling and analysis*.

## Data Availability

The data used in this data descriptor is publicly available in: https://physionet.org/content/mbrset/1.0/
All the codes used in this paper for the dataset setup, data analysis, and experiments are found in a GitHub repository at https://github.com/luisnakayama/mBRSET.

## Code Availability

All the codes used in this paper for the dataset setup, data analysis, and experiments are found in a GitHub repository at https://github.com/luisnakayama/mBRSET. When analyzing the data, best practice guidelines should be followed, and we incentivize sharing codes and models to promote reproducibility.

## Acknowledgments

The authors declare no additional acknowledgments.

## Author contributions

These authors contributed equally: Chenwei Wu and David Restrepo. Chenwei Wu, David Restrepo, and Luis Filipe Nakayama were responsible for the study design. The software development was carried out by Chenwei Wu, David Restrepo, Luis Filipe Nakayama, and Zitao Shuai. Chenwei Wu and Zitao Shuai conducted the experiments. Data collection was performed by Luis Filipe Nakayama, Nathan Santos Barboza, Maria Luiza Vieira Sousa, Raul Dias Fitterman, Alexandre Durao Alves Pereira, Caio Vinicius Saito Regatieri, Jose Augusto Stuchi, Fernando Korn Malerbi, and Rafael E. Andrade. All authors contributed to writing and editing the manuscript. Luis Filipe Nakayama is the corresponding author.

